# Quantifying the impact of the COVID-19 pandemic on invasive bacterial diseases across 27 countries and territories: prospective surveillance by the IRIS Consortium

**DOI:** 10.1101/2025.07.10.25331197

**Authors:** David Shaw, Raquel Abad, Samanta CG Almeida, Zahin Amin-Chowdhury, Adriana Bautista, Désirée Bennett, Declan Bradley, Karen Broughton, Carlo Casanova, Eun Hwa Choi, Heike Claus, Mary Corcoran, Simon Cottrell, Robert Cunney, Lize Cuypers, Tine Dalby, Heather Davies, Linda de Gouveia, Ala-Eddine Deghmane, Stefanie Desmet, Emma Dickson, Mirian Domenech, Richard Drew, Carolina Duarte, Mignon du Plessis, Helga Erlendsdóttir, Kurt Fuursted, Alyssa Golden, Desiree Henares, Birgitta Henriques-Normark, Markus Hilty, Steen Hoffmann, Michal Honskus, Hilary Humphreys, Susanne Jacobsson, Christopher Johnson, Keith A Jolley, Aníbal Kawabata, Nuala Kealy, Jana Kozakova, Karl G Kristinsson, Pavla Krizova, Alicja Kuch, Shamez N Ladhani, Thiên-Trí Lâm, María Eugenia León Ayala, Laura Lindholm, David Litt, Martin CJ Maiden, Irene Martin, Delphine Martiny, Wesley Mattheus, Noel D McCarthy, Mary Meehan, Susan Meiring, Kartyk Moganeradj, Paula Mölling, Eva Morfeldt, Julie Morgan, Robert Mulhall, Carmen Muñoz-Almagro, David Murdoch, Joy Murphy, Martin Musilek, Ludmila Novakova, Shahin Oftadeh, Zuzana Okonji, Amaresh Perez-Argüello, María Dolores Pérez-Vázquez, Monique Perrin, Malorie Perry, Benoit Prevost, Julia Villalba Primitz, Una Ren, Maria Roberts, Assaf Rokney, Merav Ron, Olga Marina Sanabria, Kevin J Scott, Julio Sempere, Lotta Siira, Ana Paula Silva de Lemos, Vitali Sintchenko, Anna Skoczyńska, Monica Sloan, Hans-Christian Slotved, Andrew J Smith, Anneke Steens, Muhamed-Kheir Taha, Maija Toropainen, Georgina Tzanakaki, Anni Vainio, Mark PG van der Linden, Nina M van Sorge, Emmanuelle Varon, Julio Vazquez, Sandra Vohrnova, Anne von Gottberg, Jose Yuste, Rosemeire Zanella, Angela B Brueggemann

**Affiliations:** University of Oxford

**Author notes:** **Corresponding author:** Prof Angela Brueggemann, Nuffield Department of Population Health, University of Oxford, United Kingdom.

## Abstract

**Background:** *Streptococcus pneumoniae, Haemophilus influenzae* and *Neisseria meningitidis* are leading causes of invasive bacterial disease worldwide. The aims of this study were to measure post-COVID-19 pandemic changes in the incidence of disease caused by these bacterial pathogens and assess the evidence for any age- or serotype/group-specific changes.

**Methods:** Prospectively, cases of invasive disease from 2018-2023 were submitted to national and/or regional microbiology reference laboratories in each of 27 countries/territories. An effective pandemic period was estimated, and interrupted time series analyses quantified the effect of the introduction and withdrawal of pandemic containment measures. Disease incidence rates for *S pneumoniae, H influenzae* and *N meningitidis* were calculated, stratified by age and serotype/group. *Streptococcus agalactiae* was investigated as a comparator invasive pathogen not transmitted via the respiratory route.

**Findings:** The median duration of pandemic containment measures among all 27 countries was 22 months (range, 14-26 months). Withdrawal of containment measures led to significant increases in invasive disease for *S. pneumoniae* (42%, incidence rate ratio [IRR] 1.42, 1.30-1.55) and *H influenzae* (68%, IRR 1.68, 1.47-1.91), but not *N meningitidis* (15%, IRR 1.15, 0.92-1.44). In the post-pandemic period, the risk of *H influenzae* disease (risk ratio [RR] 1.02, 0.91-1.14) returned to the pre-pandemic risk. Conversely, for *S pneumoniae* (RR 0.91, 0.85-0.97) and *N meningitidis* (RR 0.61, 0.54-0.69) the risk of invasive disease was significantly below pre-pandemic risk. There was no evidence for changed rates of *S. agalactiae* disease. The distribution of invasive disease by age was broadly as expected, and age-stratified disease incidence by serotype/group was similar to pre-pandemic rates, but with some notable changes.

**Interpretation:** By 2023, while the risk of invasive disease caused by *H influenzae* returned to pre-pandemic levels, the risk of *S pneumoniae* and *N meningitidis* disease remained below pre-pandemic levels. Changes were observed in the post-pandemic incidence and age-stratified distributions of some serotypes/groups.

**Research in context:** *Evidence before this study:* We searched PubMed, bioRxiv, and medRxiv for articles published up to 31 Dec 2019 (before the COVID-19 pandemic) that reported on the effects of containment measures implemented in response to a pandemic. We identified 262 papers by searching for ‘pandemic’ AND ‘microbial transmission’ OR ‘transmission’ AND ‘containment’ but none of these described the effects of implementing large-scale containment measures during a pandemic. The Invasive Respiratory Infection Surveillance (IRIS) Consortium previously reported that the incidence of invasive bacterial disease due to *S pneumoniae, H influenzae* and *N meningitidis* was significantly reduced when COVID-19 pandemic containment measures were implemented to control the transmission of SARS-CoV-2.

*Added value of this study:* We extended the previous analyses to estimate the effective containment measure period within each of 27 countries across six continents, and calculated rates of invasive bacterial disease after the pandemic containment measures were withdrawn. This demonstrated that invasive bacterial disease caused by all three pathogens increased once pandemic containment measures were removed and incidence rates exceeded the rates observed before the pandemic. Subsequently, disease caused by *H influenzae* generally returned to pre-pandemic rates, whilst disease caused by *S pneumoniae* and *N meningitidis* remained below pre-pandemic rates. Some serotype/group and age-specific changes were also observed for each of the pathogens.

*Implications of all the available evidence:* The COVID-19 pandemic had profound direct and indirect effects on global public health. The IRIS Consortium has shown that invasive bacterial disease caused by *S pneumoniae, H influenzae* and *N meningitidis* was also altered, and that changes in the circulating serotypes/groups were observed among countries participating in the IRIS Consortium. Whether or not these epidemiological changes persist, and to what extent they might affect bacterial vaccination coverage and disease rates, will be revealed over time and thus invasive disease due to these pathogens needs to be closely monitored.

## Introduction

The Invasive Respiratory Infection Surveillance (IRIS) Consortium was established early in the COVID-19 pandemic with the aim of prospectively tracking changes in the incidence and distribution of invasive bacterial disease caused by four organisms, *Streptococcus pneumoniae, Haemophilus influenzae, Neisseria meningitidis* and *Streptococcus agalactiae*. These four pathogens are leading causes of global morbidity and mortality, particularly among young children and older adults. ^1–4^

*S pneumoniae, H influenzae* and *N meningitidis* reside in the nasopharynx or throat and are transmitted through respiratory secretions, whereas *S agalactiae* is found in the healthy lower genital and gastrointestinal tract. National and regional microbiology reference laboratories participating in the IRIS Consortium actively collect isolates and provenance data for one or more of these pathogens and the laboratories have contributed data from 2018 onwards to the IRIS surveillance programme.^1–2^

A significant decrease in the incidence of invasive disease caused by *S pneumoniae, H influenzae* and *N meningitidis* was documented during the first five months of the COVID-19 pandemic.^1^ The observed reduction in invasive bacterial disease was associated with the implementation of containment measures to control the spread of SARS-CoV-2: *S pneumoniae* (incident rate ratio [IRR], 95% confidence interval [CI]) IRR 0.63, 0.57-0.70, *H influenzae* IRR 0.78, 0.62-0.99, and *N meningitidis* IRR 0.58, 0.43-0.77). No significant changes were observed for *S agalactiae* (IRR 0.90, 0.69-1.16). ^1^

This reduction in invasive bacterial disease early in the pandemic was sustained through to the end of 2021: *S pneumoniae* (risk ratio [RR], 95% CI) 0.47, 0.40-0.55, *H influenzae* 0.51, 0.40-0.66 and *N meningitidis* 0.26, 0.21-0.31. Importantly, when the cases of bacterial disease were stratified by the serotype/serogroup of the microbe or age of the patient, the distributions remained similar to what was observed before the pandemic. ^2^

In this study, the effective pandemic period for each country or region was estimated, and an interrupted time series approach was used to quantify changes in the incidence of invasive bacterial disease after the removal of COVID-19 containment measures. Age- and serotype/group-specific incidence rates of invasive bacterial disease were calculated for each study year, 2018-2023. The incidence of invasive disease in 2023 was compared to incidence rates in previous years, and significant post-pandemic changes in disease by age and serotype/group were investigated.

## Methods

### Invasive bacterial disease data collection

No patient-identifiable data were collected. National and/or regional microbiology reference laboratories in each of the participating countries/territories submitted data for cases of invasive bacterial disease caused by one or more of the four organisms under study, among all age groups, between 1 January 2018 and 31 December 2023. These data were uploaded to private projects within PubMLST (https://pubmlst.org/), and quality control checks were performed. Data were stratified by country and organism, and grouped by the month of each year.

### Estimating the COVID-19 pandemic containment period for each country

An effective pandemic period was estimated using two sets of publicly available data (appendix p1-6). First, the Oxford Blavatnik COVID-19 Government Response Tracker (OxCGRT), specifically: i) the country-specific stringency index; ii) the percentage of the population vaccinated against SARS-CoV-2; and iii) the COVID-19 cumulative case count.^5^ Second, the Google COVID-19 Community Mobility Reports (CCMR) data, specifically: i) time spent in the workplace, percentage change from baseline; and ii) time spent at residences, percentage change from baseline.^6^

The statistical technique known as change point detection was used to determine the date when the mean or variance (or both) of a time series changed significantly. This approach was applied separately to determine the country-specific date of containment measure implementation and withdrawal. The R package ‘changepoint’ was used, which employed the At Most One Change (AMOC) algorithm and the modified Bayesian Information Criterion (mBIC) penalty function to detect the position of the single most significant change in the mean and variance of the time series under analysis (appendix pp 6-7).^7^

To determine containment measure implementation dates, change point detection was applied to a slice of the country-specific CCMR workplace and residential time series from 15 February to 1 June 2020. The same approach was performed on the country-specific OxCGRT stringency index time series, analysing the time period between 1 January and 1 June 2020, and the resulting dates were compared. CCMR data were not available for Iceland (privacy concerns due to the small population).

The date when COVID-19 cases increased significantly was determined using the country-specific cumulative COVID-19 case counts, with the rationale that an increase in COVID-19 cases could be used as a proxy measure for the time point when pandemic containment measures were withdrawn and SARS-CoV-2 transmission within the human population increased significantly.^5^

### Sensitivity analyses

The first recording of national lockdown in the OxCGRT stringency index was tested as an alternative method for determining the date of introduction of containment measures. The proportion of the population vaccinated against COVID-19 was used as an alternative metric to estimate when the pandemic had ended, using a threshold value of 70% (appendix, pp 1-6).^8^ Poisson and negative binomial models were compared to assume different distributions of the data. Different approaches were also compared to capture seasonality, using month as a factor variable (time stratified model) and Fourier terms (appendix p 10).

### Calculation of disease incidence rates and population coverage

Published national census results and United Nations population estimates and projections were used to collect data on the population of each country/territory between 2018 and 2023, stratified by age group.^9–13^ Since South Korea only provides paediatric cases of invasive pneumococcal disease to IRIS, only the population aged 0 to 18 years was used in these analyses. Population estimates were used to calculate crude and serotype/group-specific invasive bacterial disease incidence rates, stratified by age group, and as offsets in the various time series models.

Population surveillance coverage data were requested from each of the microbiology reference laboratories and these data were used to adjust the invasive disease case counts to provide a more accurate estimate of the true incidence of disease (appendix p 7-8). Note that *S agalactiae* was excluded from these analyses due to low (65% or less) but consistent population coverage, and most of the laboratories collecting *S agalactiae* do not routinely serotype the isolates so serotype-specific analyses were not performed.

Several countries were excluded from age-specific analyses: Israel and Sweden did not provide age data to IRIS for *S pneumoniae* and *H influenzae* cases; Brazil only provided meningitis case counts for *S pneumoniae, H influenzae* and *N meningitidis;* South Korea only provided paediatric *S pneumoniae* case counts; Finland did not provide age data for *N meningitidis* cases; and Switzerland opted out of age-specific analyses.

### Interrupted time series analyses

Interrupted time series analyses were used to quantify the effect of containment measure introduction and withdrawal, and to estimate the relative risk of disease during and after the pandemic containment period. A segmented regression approach was employed, using generalised linear models of the quasi-Poisson family, to take account of data overdispersion (variance greater than the mean). The country population totals were used as offsets in the models. Seasonality was modelled using Fourier terms (four harmonic terms) with periods of 6 and 12 months (appendix pp 8-9). Adjusted case counts were grouped by calendar month.

Two models were fitted to each country, one that assessed the effect of containment measure introduction and another that assessed the effect of containment measure withdrawal, which yielded IRRs with 95% confidence intervals. The IRRs were calculated by exponentiating the natural logarithmic estimate of the model’s intervention dummy variable, coded ‘0’ before the intervention and ‘1’ from the time of intervention change (implementation or withdrawal). These models were used to generate counterfactuals that forecasted the assumed trend in the absence of the intervention. Observed and counterfactual case counts were compared and yielded an RR with 95% confidence intervals. RRs were obtained by dividing the observed case count by the estimated counterfactual cases count during a specific period of time (either during the pandemic or after withdrawal of containment measures). The overall results were pooled using both inverse-variance weighted and random effects meta-analytic models for comparison, treating each country as a separate study.

### Logistic regression modelling

Multivariable logistic regression models with floating absolute odds were used to determine which serotypes/groups had significantly changed in the post-pandemic period as compared to the pre-pandemic period, controlling for country/territory.^14,15^ This approach obtains confidence intervals that are based on the floated variances of the log odds ratios for the groups that are being compared with each other, allowing for more reliable estimates without relying on a baseline group. This method allows researchers to select their own reference category for comparisons.

High-prevalence serotypes/groups for each organism were chosen as the comparator: *S pneumoniae*, serotype 3; nontypable *H influenzae* (NTHi); and *N meningitidis* serogroup B. All serotypes/groups were included in the models, although for *S pneumoniae* and *N meningitidis* the serotypes/groups associated with 99% of the reported cases of invasive disease were each analysed individually. The remaining serotypes/groups were pooled together and analysed as one group called ‘other’ to improve statistical power. Bonferroni corrections were made for multiple testing of serotypes/groups within each logistic regression model. Complete case analysis was employed.

## Results

### Invasive disease cases and estimating the pandemic period

Twenty-seven national and regional microbiology reference laboratories submitted data from 2018-2023 for one or more bacterial species. In total, the number of cases of invasive disease were as follows (number of countries submitting data): 150,250 *S pneumoniae* (n=27); 19,688 *H influenzae* (n=22); 8,670 *N meningitidis* (n=19); and 11,338 *S agalactiae* (n=9). After adjusting for population coverage, a total of 230,954 cases were available for analyses.

An effective pandemic period was estimated for each country/territory, and the median duration of pandemic containment measures was 22 months (interquartile range, 21-23 months; Figure 1). The longest duration of containment measures (24-26 months) was observed in Australia, New Zealand, and South Korea, whereas the shortest duration (14-15 months) was in Brazil, Colombia, Paraguay, and South Africa (Figure 1). The sensitivity analyses produced pandemic containment measure intervals that were of a similar magnitude, and with similar start and end dates. These estimates were also broadly in agreement with other published estimates of the end of the pandemic.^8^

**Figure 1.**
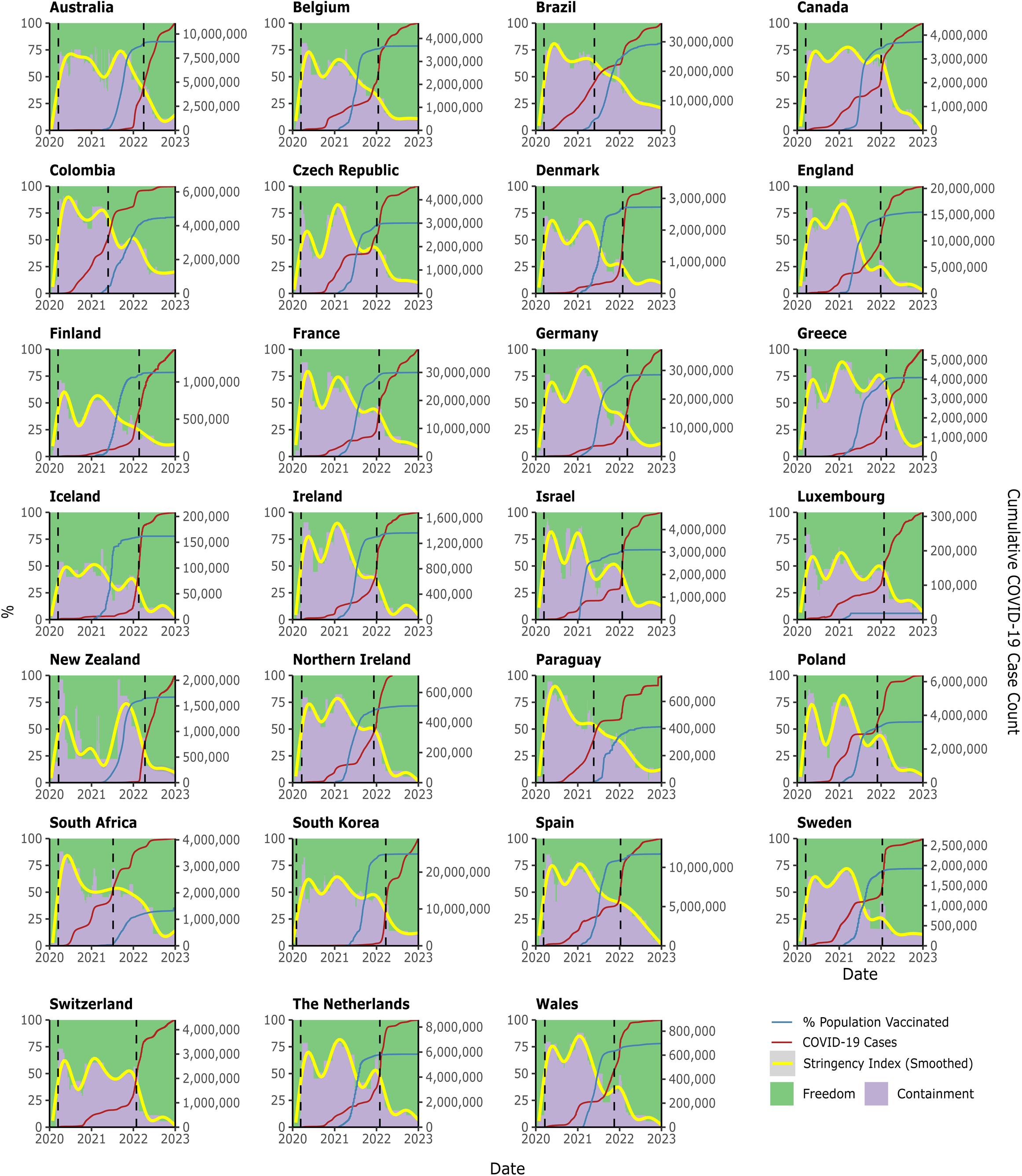
Estimated period of COVID-19 pandemic containment measures for each country/territory. The time period during which some level of containment measures was in place is marked in purple, and vertical hashed lines mark the estimated start and end dates of the pandemic period of restrictions. Note that the proportion of the population vaccinated against COVID-19 in Switzerland was not captured by the Oxford Blavatnik COVID-19 Government Response Tracker.

### Interrupted time series analyses

Pandemic containment measures were associated with a 60% reduction (RR 0.40, 0.38-0.42), 58% reduction (RR 0.42, 0.38-0.45), and 73% reduction (RR 0.27, 0.24-0.29) in the risk of *S pneumoniae*, *H influenzae* and *N meningitidis* invasive disease, respectively, during the pandemic (Figures 2 and 3). There was no significant change in the risk of *S agalactiae* invasive disease (RR 0.98, 0.89-1.07; Figure 3).

**Figure 2.**
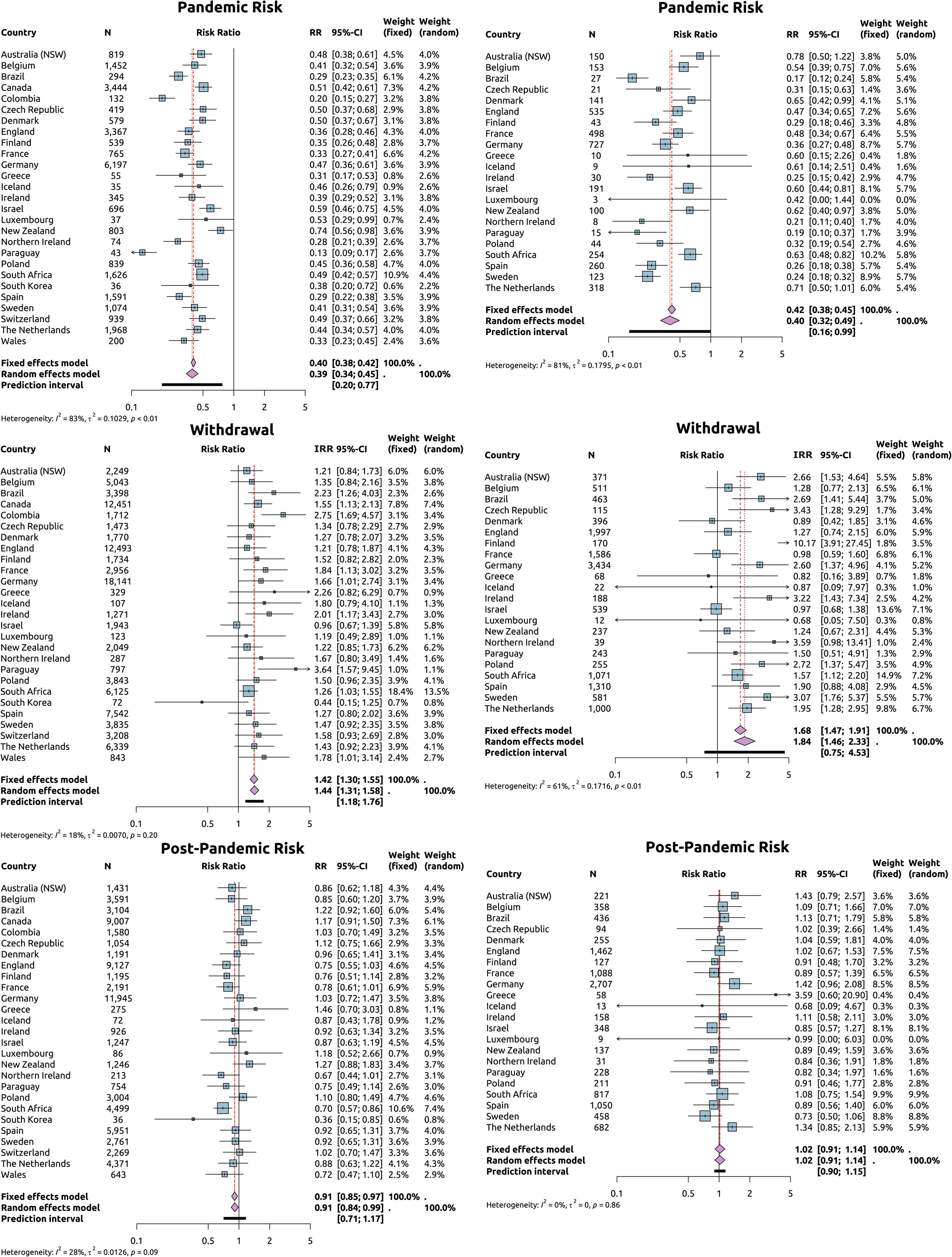
Results of the interrupted time series analyses for *S pneumoniae* (left column) and *H influenzae* (right column). ‘Pandemic risk’ refers to the risk of invasive disease during the estimated period of pandemic containment measures for each country/territory, relative to the pre-pandemic risk of invasive disease (the counterfactual of no containment measures). ‘Withdrawal’ refers to the change in rate of invasive disease after the pandemic period ended and containment measures were removed, compared to during the pandemic. (The exponentiated value of the coefficient for each variable generated an incident rate ratio.) ‘Post-pandemic risk’ refers to the risk of invasive disease in 2023 relative to the pre-pandemic risk of invasive disease (the counterfactual of no containment measures).

**Figure 3.**
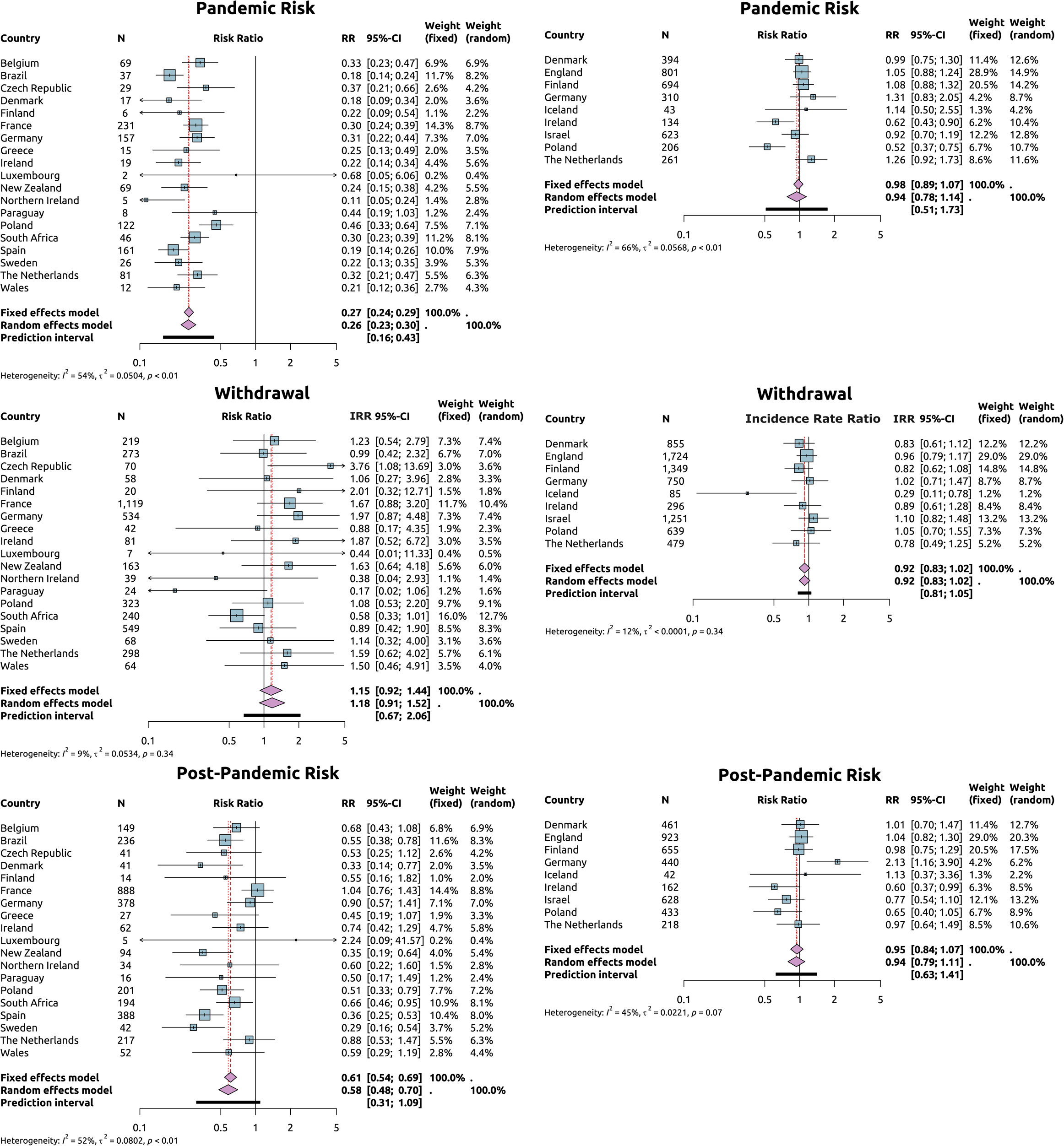
Results of the interrupted time series analyses for *N meningitidis* (left column) and *S agalactiae* (right column). ‘Pandemic risk’ refers to the risk of invasive disease during the estimated period of pandemic containment measures for each country/territory, relative to the pre-pandemic risk of invasive disease (the counterfactual of no containment measures). ‘Withdrawal’ refers to the change in rate of invasive disease after the pandemic period ended and containment measures were removed, compared to during the pandemic. ‘Post-pandemic risk’ refers to the risk of invasive disease in 2023 relative to the pre-pandemic risk of invasive disease (the counterfactual of no containment measures). Note that Iceland did not have enough *N meningitidis* cases to fit a suitable time series model.

Conversely, the withdrawal of containment measures was associated with a 42% increase (IRR 1.42, 1.30-1.55), 68% increase (IRR 1.68, 1.47-1.91), and 15% increase (IRR 1.15, 0.92-1.44) in the rate of *S pneumoniae*, *H influenzae* and *N meningitidis* invasive disease, respectively, as compared to the pandemic period (Figures 2 and 3). There was no significant change for *S agalactiae* (IRR 0.92, 0.83-1.02; Figure 3).

The post-pandemic risk of invasive disease was similar to the pre-pandemic level of risk for *H influenzae* (RR 1.02, 0.91-1.14; Figure 2). The post-pandemic risk of *S pneumoniae* invasive disease (RR 0.91, 0.85-0.97) and *N meningitidis* disease (RR 0.61, 0.54-0.69) remained below the pre-pandemic risk (Figures 2 and 3). In contrast, there was no significant change observed for *S agalactiae* (RR 0.95, 0.84-1.07; Figure 3).

### Age-stratified incidence of invasive bacterial disease between 2018 and 2023

The distribution of invasive disease across age groups was as expected in each of the six study years, that is, there was a higher incidence of disease at the two ends of the age spectrum for *S pneumoniae*, *H influenzae* and *N meningitidis*, in addition to higher *N meningitidis* disease incidence among teenagers and young adults (Figure 4). As we previously reported, there was a significant decrease in the incidence of disease during the pandemic years of 2020-2021 (as compared to the pre-pandemic years of 2018-2019) for *S pneumoniae*, *H influenzae* and *N meningitidis*.^1,2^

**Figure 4.**
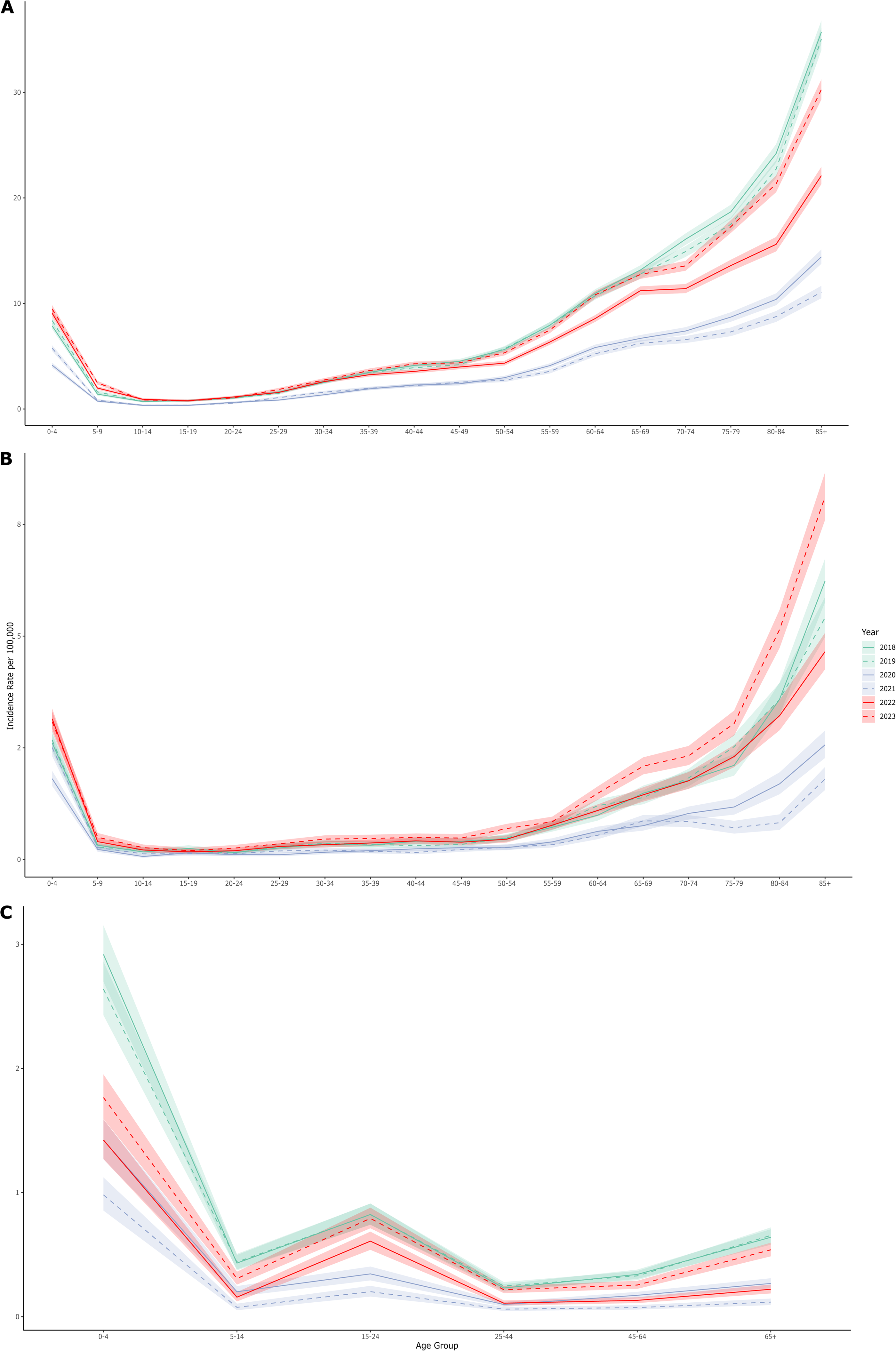
Age-stratified incidence of invasive disease caused by *S pneumoniae*, *H influenzae,* and *N meningitidis*. Overall incidence of invasive disease for each pathogen, calculated per year (2018-2023) and stratified by age group.

In contrast, as compared to the pre-pandemic years (2018-2019), the *S pneumoniae* data from 2022-2023 revealed a small increase in disease incidence among children less than 10 years of age and a return to pre-pandemic rates among older children and adults by 2023 (Figure 4). Among adults over 65 years of age in 2023, the *S pneumoniae* disease rates were approaching, but remained slightly below, pre-pandemic levels. For *H influenzae*, the incidence of invasive disease in 2023 was higher than in pre-pandemic and pandemic years across nearly all age groups and especially those adults over 50 years of age (Figure 4). Conversely, in 2023 the incidence of *N meningitidis* disease remained lower than pre-pandemic rates among children less than five years of age but approached pre-pandemic levels among all other age groups.

### Estimating relative post- vs pre-pandemic incidence of invasive disease by serotype

The post-pandemic odds of invasive disease (odds ratio [OR], 95% CI) caused by *S pneumoniae* serotype 4 (OR 1.71, 1.61-1.81), serotype 5 (OR 2.32, 1.61-3.33), serotype 9V (OR 1.69, 1.50-1.89), *H influenzae* serotype a (OR 1.29, 1.10-1.51), and *N meningitidis* serogroup Y (OR 1.23, 1.09-1.37) significantly increased as compared to the pre-pandemic years, relative to the most prevalent serotype/group for each organism and after controlling for country/region (Figure 5). Previously, we reported a small but statistically significant increase in *H influenzae* serotype b (Hib) disease in 2021^2^, but Hib disease returned to pre-pandemic levels by 2023.

**Figure 5.**
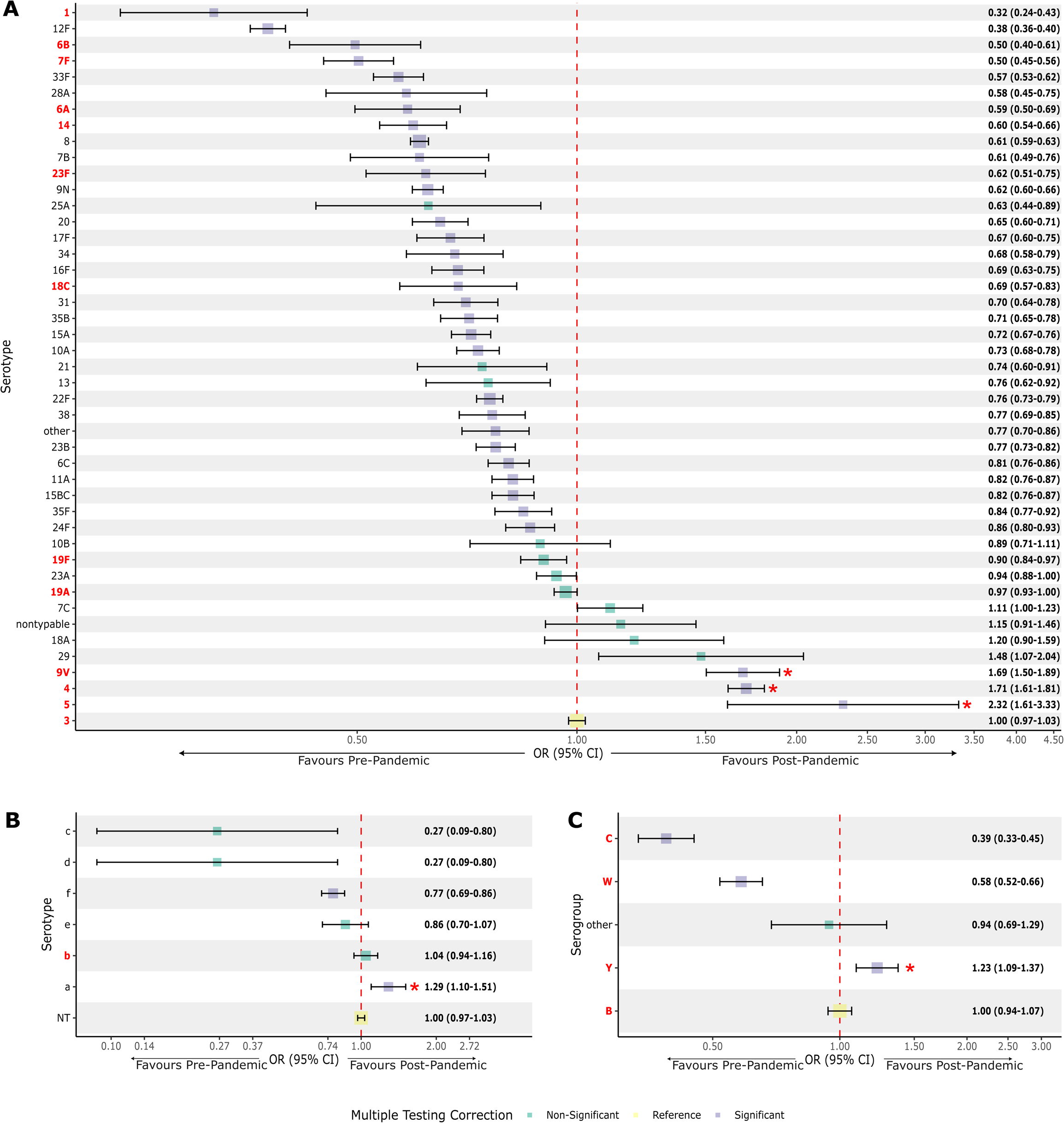
Estimated odds of invasive disease stratified by serotype or serogroup for *S pneumoniae* (A), *H influenzae* (B), and *N meningitidis* (C). Odds ratios compared the numbers of cases of invasive disease observed post-pandemic to that observed pre-pandemic, controlled for country/territory and corrected for multiple testing. For each pathogen, the reference serotype/group to which all other serotypes/groups are compared is marked in yellow. The 13-valent pneumococcal conjugate vaccine serotypes/groups are denoted by red font. Floated variances around the baseline category allow the reader to calculate their own comparison group. A red asterisk denotes a statistically significant increase in the odds of a particular serotype/group in the post-pandemic period, after correction for multiple testing.

### Age-stratified incidence of invasive bacterial disease by serotype or serogroup

The distribution of *S pneumoniae* invasive disease across the age spectrum for each of 30 serotypes included in one or more of the pneumococcal conjugate vaccines (PCVs) was broadly as expected, that is, the incidence of disease was highest in the youngest and oldest age groups (Figure 6). Five serotypes revealed an unusual pattern of disease across the age spectrum: serotype 4 (increased in patients 40+ years of age; higher incidence in 2023 compared to previous years); serotype 7F (similar incidence among patients 30+ years irrespective of age; lower incidence in 2023 compared to previous years among older individuals); serotype 8 (steady increase in the incidence of disease from 10+ years of age; lower incidence in 2023 than previous years); serotype 9V (increased disease across all but the 80+ age groups in 2023); and serotype 12F (relatively similar incidence rates among patients 35+ years of age; lower incidence in 2023 than previous years).

**Figure 6.**
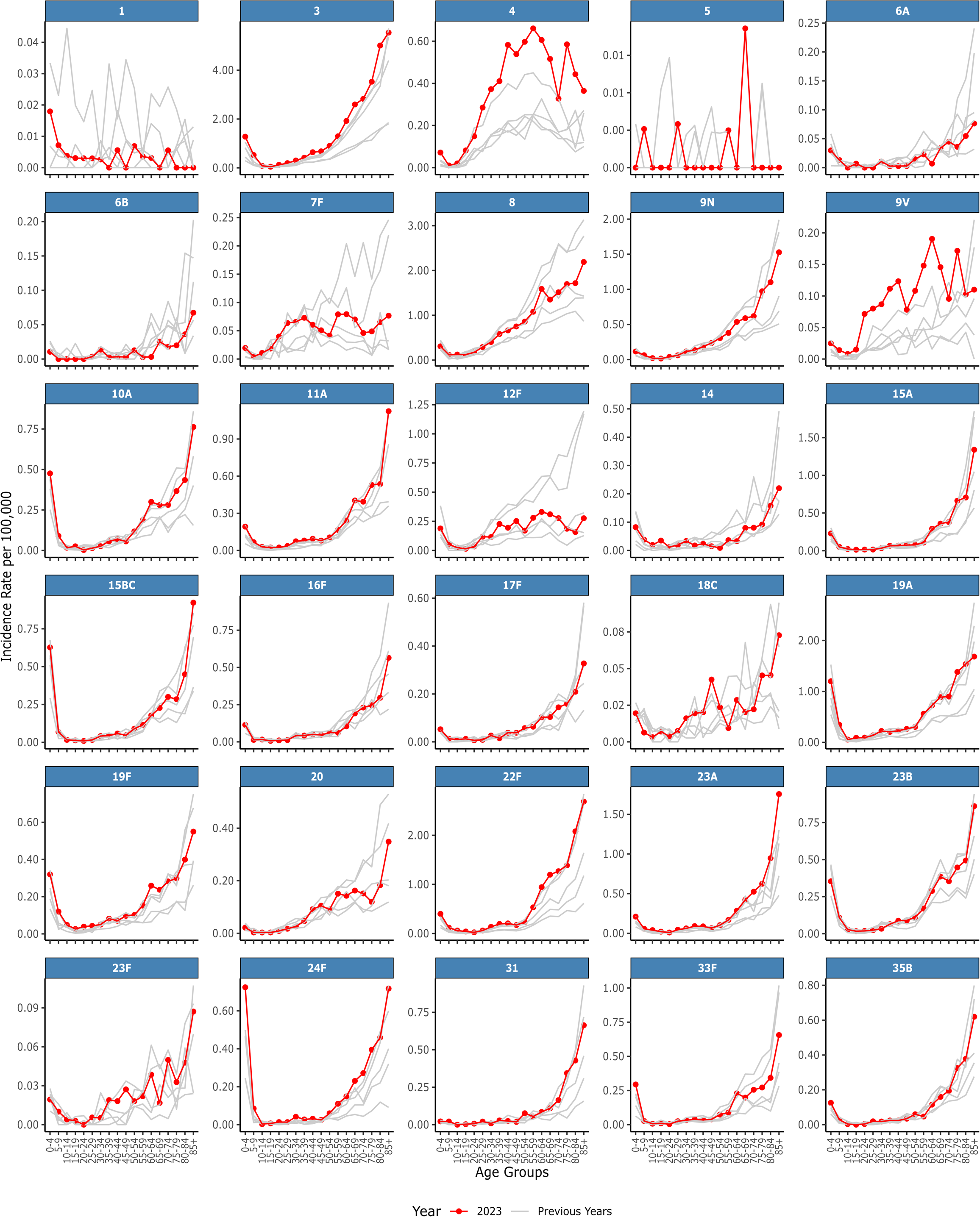
Age-specific incidence rates of invasive disease caused by *S pneumoniae* serotypes. Incidence rates were calculated for each of the 30 serotypes included in any pneumococcal conjugate vaccine, stratified by age group. The red line indicates 2023 incidence rates, and the grey lines display the incidence rates for each of the previous years from 2018-2022.

The remaining 25 serotypes included in any PCV demonstrated similar or lower incidence rates of invasive disease in 2023 relative to previous years, and the age distributions were as expected: serotypes 3, 6A, 6B, 9N, 10A, 11A, 14, 15A, 15BC, 16F, 17F, 18C, 19A, 19F, 20, 22F, 23A, 23B, 23F, 24F, 31, 33F, and 35B (Figure 6). Serotypes 1 and 5 were only infrequently a cause of disease in any of the study years. Overall, in 2023, the incidence of *S pneumoniae* invasive disease among children less than five years of age was highest for serotype 3 (1.28 cases per 100,000) and serotype 19A (1.20 cases per 100,000). Among the adults 65 to 85+ years of age, *S pneumoniae* invasive disease was highest for serotype 3 (range, 2.60-5.54 cases per 100,000) and serotype 8 (range, 1.35-2.19 cases per 100,000).

The highest incidence of *H influenzae* invasive disease was observed for NTHi and the age distribution of NTHi cases in 2023 was as expected, compared to previous years (Figure 7). The incidence rate among children less than five years of age in 2023 was 1.70 cases per 100,000 but ranged from 1.67-7.31 cases per 100,000 among adults 65 to 85+ years of age. The incidence of invasive disease caused by *H influenzae* serotypes a, b, e and f was low and followed the expected age distributions for each serotype. Hib disease was rare among all age groups apart from children less than 5 years of age, but even in this youngest age group the incidence rate was fewer than 1 case per 100,000.

**Figure 7.**
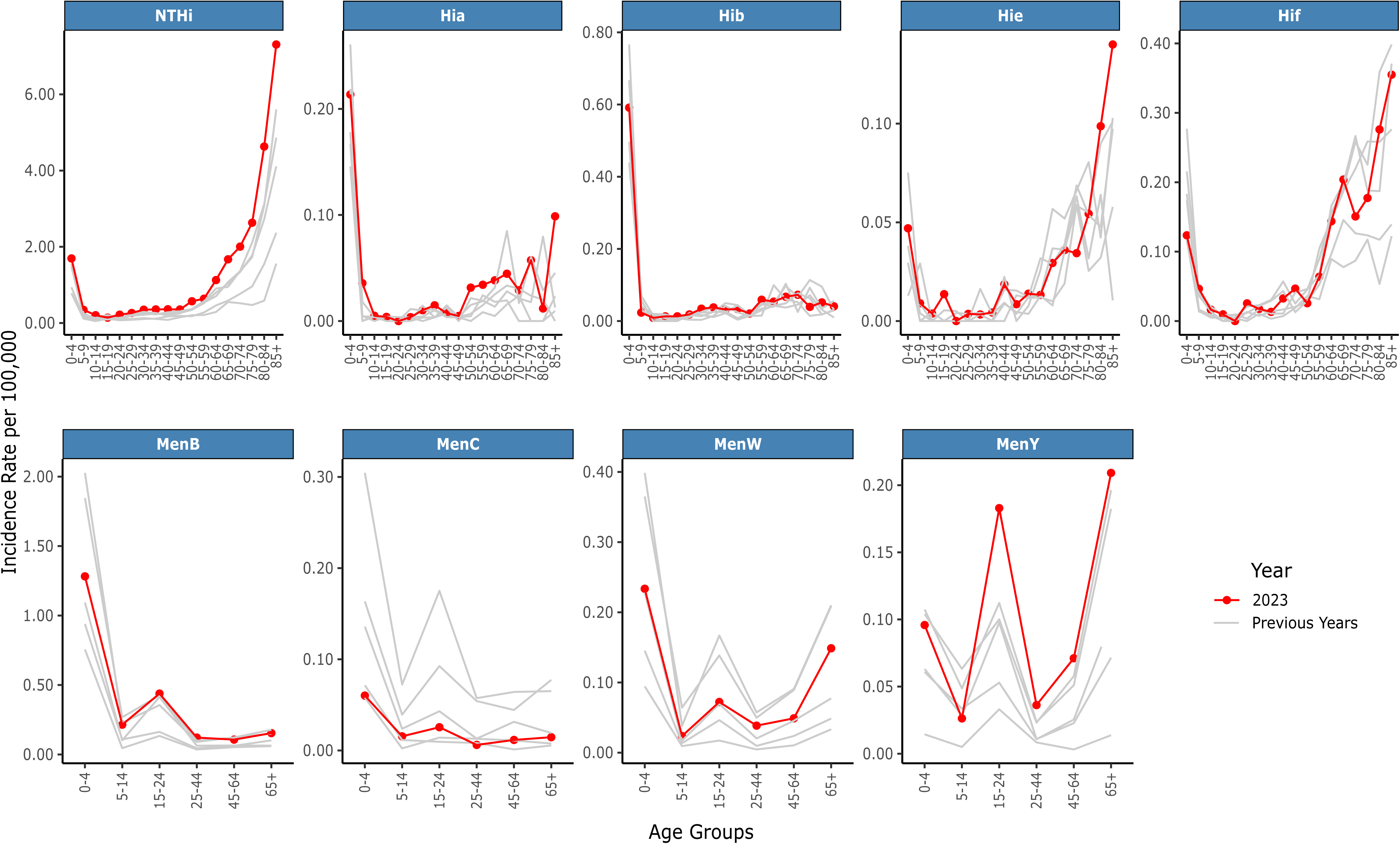
Age-specific incidence rates of invasive disease caused by *H influenzae* (top row) and *N meningitidis* (bottom row). Incidence rates were calculated for five *H influenzae* serotypes (serotypes c and d were rare) and four *N meningitidis* serogroups (serogroup A was rare), stratified by age group. NTHi, nontypable *H influenzae*; Hia-Hif, *H influenzae* serotypes a to f, respectively; MenB, C, W, Y, *N meningitidis* serogroup B, C, W, Y, respectively. The red line indicates 2023 incidence rates, and the grey lines display the incidence rates for each of the previous years from 2018-2022.

Finally, *N meningitidis* invasive disease caused by either serogroup A or C was rare, and the incidence of serogroup C disease was lower in 2023 than in previous years (Figure 7). Invasive disease due to serogroups B and W were as expected or lower in 2023 compared to previous years and the age distribution of cases was as expected. The incidence of serogroup Y disease in 2023 was higher than in previous years, although remained very low overall (peak incidence 0.21 cases per 100,000 in the 65+ age group).

## Discussion

This study provides important insights into the burden of invasive bacterial disease in the post-COVID-19 pandemic period across 27 countries/territories. The main finding was that the incidence of invasive disease caused by *S pneumoniae* and *H influenzae* temporarily increased above pre-pandemic levels after the COVID-19 containment measures were withdrawn, but *H influenzae* returned to the pre-pandemic risk of disease by 2023 and *S pneumoniae* nearly did so.

The changes could be due to an increase in the number of susceptible individuals during the pandemic because the containment measures led to decreased transmission of, and exposure to, these pathogens. This is sometimes referred to as ‘immunity debt’.^16^ Disruptions to routine vaccination programmes during the pandemic may also account for some of the increased risk of invasive disease, particularly in children.^17–20^ There is also a high incidence of disease among older age groups, combined with relatively low vaccine uptake.^21^

The post-pandemic period risk of invasive disease caused by *N meningitidis* was slightly different: there was a small temporary increase in *N meningitidis* disease after COVID-19 containment measures were withdrawn, but subsequently the risk of disease again dropped below pre-pandemic levels, although not to the same level of reduced risk as during the pandemic when containment measures were in force. The explanation for changes in the risk of *N meningitidis* disease is most likely similar to *S pneumoniae* and *H influenzae* (decreased transmission and exposure) and changes in vaccination programmes. There were no significant changes in invasive disease caused by *S agalactiae* at any point during the study period.

Importantly, this study revealed shifts in the serotypes/groups causing disease post-pandemic. For example, among *S pneumoniae*, there was a significant increase in three vaccine serotypes (4, 5, and 9V, although the incidence of serotype 5 disease was low). Also among *H influenzae* and *N meningitidis* there was a significant increase in serotype a and serogroup Y, respectively, although the incidence of disease was low overall. Notably, the age distribution of disease caused by five *S pneumoniae* vaccine serotypes (4, 7F, 8, 9V and 12F) was also different from what was expected based on pre-pandemic data.

One explanation for these findings is that observed changes are due to random variation, as bacterial populations emerge from the bottleneck created by the containment measures. Such changes may persist or return to patterns that are similar to those observed before the pandemic. For example, we previously reported a significant increase in Hib infections,^2^ but Hib disease has now returned to pre-pandemic levels. Other observed changes in disease patterns may persist as the bacterial population restructures after the pandemic. The outcome will be revealed in time and these findings reinforce the necessity of active surveillance to monitor the age- and serotype/group-specific patterns of disease for these pathogens. Vaccination programmes must also be focused on achieving wide coverage for all eligible populations and be alert to possible vaccine-related changes in the patterns of disease.

Estimating the pandemic period for each country was a key component of these analyses. Data from the CCMR for time spent at home and at work provided a reliable measure of when containment measure restrictions were introduced, but determining when containment measures were removed based on the same type of data was less reliable, in part because working from home practices changed significantly since the pandemic.^1,2,6^ Instead, the SARS-CoV-2 vaccination rates and increase in COVID-19 case counts were used in this study as a proxy for the withdrawal of containment measures in each of the countries. Sensitivity analyses suggested that even if withdrawal dates varied, the overall conclusions remained consistent, which provided reassurance that the overall findings were uniform in direction even if there was some variation in magnitude.

Limitations of this study could include missing data (i.e. cases not reported to the reference laboratories) despite our best efforts to mitigate against this, and could potentially bias the estimates; however, the consistency in trends across countries suggests that the overall findings are robust and valid. The quantified effects might also be influenced by unmeasured confounders, such as preceding or co-infection with a respiratory virus (e.g. influenza), but we do not have relevant data across all countries to address this directly. Finally, the countries currently participating in IRIS are middle- to high-income countries with well-established and robust surveillance programmes, so these findings might not be generalisable to low-income countries.

The main strengths of the IRIS Consortium and these findings are that the participating laboratories are accredited microbiology reference laboratories with wide population coverage and longstanding surveillance programmes, and they provide accurate, precise, and reliable data, which minimises selection and measurement bias. Some participating laboratories capture case data from less than 100% of their total population, and thus for these analyses adjustments were made for population coverage, which improved the generalisability of the estimates presented here. Notably, the large size of the bacterial datasets and multi-national composition of the IRIS Consortium provide considerable statistical power and minimise the role of chance in these findings. Where appropriate, corrections were made for multiple testing, and this minimised false positives. Importantly, calculating age-specific rates allowed for appropriate comparisons between countries, and using population denominators enhanced the estimated incidence rates, which provided a more accurate assessment of age- and serotype/group-specific disease.

The COVID-19 pandemic had a profound effect on global health, and these findings demonstrate the indirect effects of pandemic containment measures taken to control the spread of SARS-CoV-2. The collaboration of the IRIS Consortium highlights the importance of active, high quality, sustained national surveillance in monitoring changes in the epidemiology of invasive diseases over time. The IRIS Consortium also highlights the value and impact of international data sharing. Some of the changes reported here may be transitory and not prove to be impactful to human health or vaccination programmes in the longer term; however, other changes might be, and thus it is important to be able to detect, assess, and respond to such changes for the benefit of global public health. During the pandemic, the public in most countries embraced the need for improved hand hygiene and respiratory etiquette, which contributed to infection prevention and control measures. There is a case for public health messaging to continue to advocate for these personal hygiene measures to reduce the transmission of microbes in general, not only during pandemics.

### Data statement

Data was stored in private projects within the BIGSdb platform of PubMLST. Data analyses were performed with R software v4 in RStudio. The Oxford Biomedical Research Computing (BMRC) high performance computers were used for computationally intensive data analyses. Analysis code is available on GitHub (https://github.com/brueggemann-lab).

### Data sharing

It is not possible to share the study data because doing so would risk identifying individual cases of invasive disease in countries with small numbers of cases.

## Supporting information

SuppFig_1

## Authors’ contributions

ABB, MPGvdL, KAJ and MCJM conceived the idea for the IRIS Consortium, set up the database infrastructure, and recruited laboratories to join IRIS. All authors were involved in the acquisition, processing, and/or validation of microbiological data, and/or Oxford COVID-19 Government Response Tracker (OxCGRT) data. Three authors (ABB, DS and KAJ) directly accessed, curated, and verified the underlying data reported in the manuscript. DS and ABB analysed and interpreted the data. DS created the figures. DS and ABB wrote the first draft of the paper. All authors reviewed and critically revised the paper for important intellectual content and approved the final version to be published.

## Acknowledgments

We are grateful to the many hospitals and clinical microbiology laboratories that have submitted data and isolates to the reference laboratories participating in the IRIS Consortium. We are also grateful to all laboratory personnel who made important contributions to the microbiological data analysed in this study, including: Minna Haanpää, Kirsi Mäkisalo and Elina Yamazaki (Finland) for their contribution in the laboratory; Trang Nguyen (Australia) for providing invasive *H influenzae* typing data; and Yeison Torres (Colombia) for assistance with the Colombian data.

## Authors by country

**Australia**

Shahin Oftadeh
Vitali Sintchenko

**Belgium**

Benoit Prevost
Delphine Martiny
Lize Cuypers
Stefanie Desmet
Wesley Mattheus

**Brazil**

Ana Paula Silva de Lemos
Rosemeire Zanella
Samanta Almeida

**Canada**

Alyssa Golden
Irene Martin

**Colombia**

Adriana Bautista
Carolina Duarte
Olga Marina Sanabria

**Czech Republic**

Jana Kozakova
Ludmila Novakova
Martin Musilek
Michal Honskus
Pavla Krizova
Sandra Vohrnova
Zuzana Okonji

**Denmark**

Hans-Christian Slotved
Kurt Fuursted
Steen Hoffmann
Tine Dalby

**England**

Angela Brueggemann
David Litt
David Shaw
Karen Broughton
Kartyk Moganeradj
Keith Jolley
Martin Maiden
Shamez Ladhani
Zahin Amin-Chowdhury

**Finland**

Anni Vainio
Laura Lindholm
Lotta Siira
Maija Toropainen

**France**

Ala-Eddine Deghmane
Emmanuelle Varon
Muhamed-Kheir Taha

**Germany**

Heike Claus
Mark van der Linden
Thiên-Trí Lâm

**Greece**

Georgina Tzanakaki

**Iceland**

Helga Erlendsdottir
Karl Kristinsson

**Ireland**

Désirée Bennett
Hilary Humphreys
Julia Villalba Primitz
Mary Corcoran
Mary Meehan
Noel McCarthy
Nuala Kealy
Richard Drew
Robert Cunney
Robert Mulhall

**Israel**

Assaf Rokney
Merav Ron

**Luxembourg**

Monique Perrin

**New Zealand**

David Murdoch
Heather Davies
Julie Morgan
Una Ren

**Northern Ireland**

Declan Bradley
Emma Dickson
Joy Murphy
Monica Sloan

**Paraguay**

Anibal Kawabata
María Eugenia León Ayala

**Poland**

Alicja Kuch
Anna Skoczyńska

**Scotland**

Andrew Smith
Kevin Scott

**South Africa**

Anne von Gottberg
Linda de Gouveia
Mignon du Plessis
Susan Meiring

**South Korea**

Eun Hwa Choi

**Spain**

Amaresh Perez-Argüello
Carmen Muñoz-Almagro
Desiree Henares
Jose Yuste
Julio Sempere
Julio Vazquez
María Dolores Pérez-Vázquez
Mirian Domenech
Raquel Abad

**Sweden**

Birgitta Henriques-Normark
Eva Morfeldt
Paula Mölling
Susanne Jacobsson

**Switzerland**

Carlo Casanova
Markus Hilty

**The Netherlands**

Anneke Steens
Nina van Sorge

**Wales**

Christopher Johnson
Malorie Perry
Maria Roberts
Simon Cottrell

## References

1. Brueggemann AB, Jansen van Rensburg MJ, Shaw D, et al. Changes in the incidence of invasive disease due to *Streptococcus pneumoniae*, *Haemophilus influenzae*, and *Neisseria meningitidis* during the COVID-19 pandemic in 26 countries and territories in the Invasive Respiratory Infection Surveillance Initiative: a prospective analysis of surveillance data. Lancet Digit Health. 2021 Jun;3(6):e360–e370. doi: 10.1016/S2589-7500(21)00077-7.

2. Shaw D, Abad R, Amin-Chowdhury Z, et al. Trends in invasive bacterial diseases during the first 2 years of the COVID-19 pandemic: analyses of prospective surveillance data from 30 countries and territories in the IRIS Consortium. Lancet Digit Health. 2023 Sep;5(9):e582–e593. doi: 10.1016/S2589-7500(23)00108-5.

3. Shaw D, Torreblanca RA, Amin-Chowdhury Z, et al. The importance of microbiology reference laboratories and adequate funding for infectious disease surveillance. Lancet Digit Health. 2025 Apr;7(4):e275–e281. doi: 10.1016/S2589-7500(24)00241-3.

4. Estimating the impact of vaccines in reducing antimicrobial resistance and antibiotic use: technical report. Geneva: World Health Organization; 2024. https://iris.who.int/

5. Hale T, Angrist N, Goldszmidt R, Kira B, Petherick A, Phillips T, Webster S, Cameron-Blake E, Hallas L, Majumdar S, Tatlow H. A global panel database of pandemic policies (Oxford COVID-19 Government Response Tracker). Nat Hum Behav. 2021 Apr;5(4):529–538. doi: 10.1038/s41562-021-01079-8.

6. https://www.google.com/covid19/mobility/

7. Killick R, Eckley IA. changepoint: An R package for changepoint analysis. Journal of statistical software. 2014 Jun 25;58:1–9.

8. Ioannidis JP. The end of the COVID-19 pandemic. European journal of clinical investigation. 2022 Jun;52(6):e13782.

9. United Nations. World Population Prospects - Population Division. url: https://population.un.org/dataportal/home?df=868bb683-3206-42f7-ab2e-a9170eccf221 (visited on 6 Sept. 2024).

10. World Bank Open Data. url: https://data.worldbank.org (visited on 6 Sept. 2024).

11. Australian Bureau of Statistics. 2021 New South Wales, Census All Persons. url: https://www.abs.gov.au/census/find-census-data/quickstats/2021/1 (visited on 6 Sept. 2024).

12. Indescat. Population and Housing Census, Catalunya. url: https://www.idescat.cat/pub/?id=censph&n=10&lang=en&hist=taules%2Fv2%2Fcensph%2F10%2F5975%2Fcat%2Fdata%3F_LAST_%3D4%26lang%3Den%5Ec%3D3%2Fr%3D2%2Ft%3D0d%2C0%3B-2c%3B-3c%2Fe%3D0 (visited on 6 Sept. 2024).

13. Office for National Statistics. Estimates of the population for the UK, England, Wales, Scotland, and Northern Ireland. url: https://www.ons.gov.uk/peoplepopulationandcommunity/populationandmigration/populationestimates/datasets/populationestimatesforukenglandandwalesscotlandandnorthernireland (visited on 6 Sept. 2024).

14. Easton DF, Peto J, Babiker AG. Floating absolute risk: an alternative to relative risk in survival and case-control analysis avoiding an arbitrary reference group. Statistics in medicine. 1991 Jul;10(7):1025–35.

15. Plummer M. Improved estimates of floating absolute risk. Statistics in medicine. 2004 Jan 15;23(1):93–104.

16. Munro AP, House T. Cycles of susceptibility: Immunity debt explains altered infectious disease dynamics post-pandemic. Clin Infect Dis. 2024 Oct 11:ciae493.

17. Causey K, Fullman N, Sorensen RJ, Galles NC, Zheng P, Aravkin A, Danovaro-Holliday MC, Martinez-Piedra R, Sodha SV, Velandia-González MP, Gacic-Dobo M. Estimating global and regional disruptions to routine childhood vaccine coverage during the COVID-19 pandemic in 2020: a modelling study. The Lancet. 2021 Aug 7;398(10299):522–34.

18. Shet A, Carr K, Danovaro-Holliday MC, Sodha SV, Prosperi C, Wunderlich J, Wonodi C, Reynolds HW, Mirza I, Gacic-Dobo M, O’Brien KL. Impact of the SARS-CoV-2 pandemic on routine immunisation services: evidence of disruption and recovery from 170 countries and territories. The Lancet Global Health. 2022 Feb 1;10(2):e186–94.

19. Basu S, Ashok G, Debroy R, Ramaiah S, Livingstone P, Anbarasu A. Impact of the COVID-19 pandemic on routine vaccine landscape: A global perspective. Human Vaccines & Immunotherapeutics. 2023 Jan 2;19(1):2199656.

20. Hartner AM, Li X, Echeverria-Londono S, Roth J, Abbas K, Auzenbergs M, de Villiers MJ, Ferrari MJ, Fraser K, Fu H, Hallett T. Estimating the health effects of COVID-19-related immunisation disruptions in 112 countries during 2020–30: a modelling study. The Lancet Global Health. 2024 Apr 1;12(4):e563–71.

21. Weinberger, B. Vaccines for the elderly: current use and future challenges. Immun Ageing 2018; 15, 3.

