## Appendix for "Quantifying the impact of the COVID-19 pandemic on invasive bacterial diseases across 27 countries and territories: prospective surveillance by the IRIS Consortium"

#### Supplementary Material

#### 1 Pandemic Time Periods

| Dataset | Project Start Date | Project End Date | Variables Selected | Notes |
|---|---|---|---|---|
| Google COVID-19 Community Mobility Reports | 15/02/2020 | 15/10/2022 | 1. Workplace percent change from baseline<br>2. Residential percent change from baseline | No data for Iceland<br>UK's devolved nations aggregated together* |
| Oxford Blavatnik COVID-19 Government Response Tracker | 01/01/2020 | 31/12/2022 | 1. Stringency index<br>2. Confirmed cases<br>3. Population vaccinated (%) | Data available for each country participating in the IRIS Consortium |

*England, Northern Ireland, Scotland, Wales
IRIS: Invasive Respiratory Infection Surveillance, UK: United Kingdom

**Supplementary Table 1**: Datasets and variables analysed.

| Country | Containment Measure | | Interval Duration | Stringency Index (%) | | | | | | | Max Duration# |
|---|---|---|---|---|---|---|---|---|---|---|---|
| | Implementation | Withdrawal | Years (Months) | Mean (SD) | Median (IQR) | Min | Max | ≥50%* (Days) | ≥75%* (Days) | Time at Max (Days) | % |
| Australia | 18/03/2020 | 01/04/2022 | 2.04 (24.4) | 61.6 (12.0) | 67.1 (52.3-71.8) | 35.3 | 78.2 | 593 | 81 | 5 | 0.7 |
| Belgium | 13/03/2020 | 16/01/2022 | 1.85 (22.1) | 55.1 (13.5) | 52.8 (47.2-63.0) | 23.1 | 81.5 | 484 | 93 | 46 | 6.8 |
| Brazil | 13/03/2020 | 26/05/2021 | 1.2 (14.4) | 69.4 (7.7) | 69.9 (63.4-74.5) | 38.0 | 81.0 | 436 | 95 | 53 | 12.1 |
| Canada | 15/03/2020 | 02/01/2022 | 1.8 (21.6) | 69.3 (5.4) | 69.9 (65.0-74.5) | 24.1 | 76.4 | 656 | 127 | 3 | 0.5 |
| Colombia | 16/03/2020 | 26/05/2021 | 1.19 (14.3) | 77.3 (10.3) | 81 (69.4-87.0) | 45.4 | 90.7 | 436 | 276 | 9 | 2.1 |
| Czech Republic | 10/03/2020 | 04/01/2022 | 1.82 (21.8) | 53.9 (16.1) | 47.2 (40.4-69.4) | 25.0 | 82.4 | 309 | 98 | 10 | 1.5 |
| Denmark | 10/03/2020 | 28/01/2022 | 1.89 (22.6) | 50.1 (15.3) | 53.4 (39.1-63.0) | 18.3 | 72.2 | 402 | 0 | 28 | 4.1 |
| England | 21/03/2020 | 28/12/2021 | 1.77 (21.3) | 58.0 (21.8) | 63.9 (32.9-74.3) | 23.1 | 88.0 | 483 | 162 | 62 | 9.6 |
| Finland | 15/03/2020 | 20/02/2022 | 1.94 (23.2) | 42.9 (12.5) | 40.7 (33.4-52.3) | 18.5 | 71.3 | 254 | 0 | 18 | 2.5 |
| France | 13/03/2020 | 24/01/2022 | 1.87 (22.4) | 57.8 (15.1) | 51.9 (44.8-70.8) | 36.3 | 88.0 | 350 | 150 | 55 | 8.1 |
| Germany | 15/03/2020 | 09/03/2022 | 1.98 (23.8) | 59.7 (16.1) | 61.1 (43.1-75.0) | 28.3 | 85.2 | 482 | 210 | 5 | 0.7 |
| Greece | 13/03/2020 | 16/02/2022 | 1.93 (23.2) | 69.9 (11.5) | 71.3 (59.7-80.6) | 40.7 | 88.9 | 693 | 231 | 17 | 2.4 |
| Iceland | 15/03/2020 | 17/02/2022 | 1.93 (23.1) | 41.5 (9.4) | 40.7 (35.8-50.0) | 17.6 | 65.7 | 181 | 0 | 19 | 2.7 |
| Ireland | 14/03/2020 | 03/01/2022 | 1.81 (21.7) | 63.3 (19.0) | 61.6 (43.2-83.3) | 39.8 | 90.7 | 422 | 250 | 42 | 6.4 |
| Israel | 12/03/2020 | 24/01/2022 | 1.87 (22.4) | 59.0 (17.6) | 53.2 (48.6-75.0) | 22.2 | 94.4 | 448 | 194 | 6 | 0.9 |
| Luxembourg | 12/03/2020 | 27/01/2022 | 1.88 (22.5) | 49.2 (11.2) | 47.2 (41.7-55.6) | 22.2 | 79.6 | 259 | 34 | 34 | 5.0 |
| New Zealand | 20/03/2020 | 12/04/2022 | 2.06 (24.7) | 44.3 (25.5) | 39.4 (22.2-65.3) | 22.2 | 96.3 | 272 | 151 | 54 | 7.2 |
| Northern Ireland | 20/03/2020 | 09/12/2021 | 1.72 (20.7) | 64.0 (11.6) | 63.9 (52.8-75.0) | 37.0 | 82.4 | 629 | 167 | 60 | 9.5 |
| Paraguay | 09/03/2020 | 20/05/2021 | 1.2 (14.4) | 69.1 (14.6) | 67.6 (55.6-81.5) | 5.6 | 94.4 | 428 | 198 | 42 | 9.6 |
| Poland | 11/03/2020 | 30/11/2021 | 1.72 (20.7) | 55.9 (18.5) | 55.9 (38.4-73.2) | 23.1 | 87.0 | 361 | 155 | 11 | 1.7 |
| South Africa | 17/03/2020 | 08/07/2021 | 1.31 (15.7) | 61.7 (15.9) | 56.5 (48.1-76.8) | 38.9 | 88.0 | 276 | 145 | 36 | 7.5 |
| South Korea | 03/02/2020 | 23/03/2022 | 2.13 (25.6) | 51.7 (9.3) | 50.5 (44.6-57.9) | 23.1 | 82.4 | 465 | 27 | 12 | 1.5 |
| Spain | 09/03/2020 | 09/01/2022 | 1.84 (22.0) | 60.6 (13.3) | 63.4 (47.7-71.3) | 25.0 | 85.2 | 454 | 72 | 35 | 5.2 |
| Sweden | 15/03/2020 | 12/01/2022 | 1.83 (21.9) | 52.1 (17.4) | 58.3 (41.4-64.8) | 15.6 | 71.1 | 473 | 0 | 3 | 0.4 |
| Switzerland | 15/03/2020 | 27/01/2022 | 1.87 (22.4) | 51.9 (9.7) | 50 (44.4-60.2) | 29.2 | 73.2 | 428 | 0 | 41 | 6.0 |
| The Netherlands | 11/03/2020 | 30/01/2022 | 1.89 (22.7) | 59.5 (14.9) | 62 (45.4-75.0) | 20.8 | 82.4 | 476 | 180 | 16 | 2.3 |
| Wales | 20/03/2020 | 15/11/2021 | 1.66 (19.9) | 62.2 (19.2) | 63 (50.0-77.8) | 23.1 | 88.0 | 504 | 200 | 59 | 9.8 |

*Time in days spent at or above stringency index value
#Percentage of total time (interval duration) spent at maximum stringency
IQR: interquartile range, SD: standard deviation

**Supplementary Table 2**: Results of change point detection, searching for mean time series changes using the Oxford Blavatnik COVID-19 Government Response Tracker (OxCGRT) stringency index and cumulative COVID-19 case count.

| Country | Containment Measure Implementation Date | |
|---|---|---|
| | Residential Time Series | Workplace Time Series |
| Australia | 22/03/2020 | 23/03/2020 |
| Belgium | 15/03/2020 | 15/03/2020 |
| Brazil | 19/03/2020 | 19/03/2020 |
| Canada | 16/03/2020 | 15/03/2020 |
| Colombia | 19/03/2020 | 19/03/2020 |
| Czech Republic | 15/03/2020 | 15/03/2020 |
| Denmark | 11/03/2020 | 11/03/2020 |
| Finland | 15/03/2020 | 16/03/2020 |
| France | 16/03/2020 | 16/03/2020 |
| Germany | 16/03/2020 | 16/03/2020 |
| Greece | 14/03/2020 | 14/03/2020 |
| Ireland | 15/03/2020 | 15/03/2020 |
| Israel | 14/03/2020 | 14/03/2020 |
| Luxembourg | 15/03/2020 | 15/03/2020 |
| New Zealand | 23/03/2020 | 23/03/2020 |
| Paraguay | 14/03/2020 | 16/03/2020 |
| Poland | 12/03/2020 | 14/03/2020 |
| South Africa | 26/03/2020 | 26/03/2020 |
| South Korea | 21/02/2020 | 21/02/2020 |
| Spain | 13/03/2020 | 14/03/2020 |
| Sweden | 15/03/2020 | 15/03/2020 |
| Switzerland | 16/03/2020 | 16/03/2020 |
| The Netherlands | 15/03/2020 | 15/03/2020 |
| United Kingdom | 22/03/2020 | 22/03/2020 |

*Implementation date occurred prior to data recording
No data available for Iceland
England, Northern Ireland, Scotland and Wales aggregated together as the United Kingdom

**Supplementary Table 3**: Containment measure implementation dates determined via change point detection, searching for changes in the mean of the residential and workplace time series from Google COVID-19 Community Mobility Reports (CCMR).

| Country | Containment Measure | | Interval Duration | Immunity Threshold* | New Duration | Difference** |
|---|---|---|---|---|---|---|
| | Implementation | Withdrawal | Years (Months) | Pandemic End date | Years (Months) | Months |
| Australia | 18/03/2020 | 01/04/2022 | 2.04 (24.4) | 21/11/2021 | 1.68 (20.1) | -4.3 |
| Belgium | 13/03/2020 | 16/01/2022 | 1.85 (22.1) | 30/08/2021 | 1.46 (17.6) | -4.6 |
| Brazil | 13/03/2020 | 26/05/2021 | 1.2 (14.4) | 05/02/2022 | 1.9 (22.8) | 8.4 |
| Canada | 15/03/2020 | 02/01/2022 | 1.8 (21.6) | 28/09/2021 | 1.54 (18.5) | -3.2 |
| Colombia | 16/03/2020 | 26/05/2021 | 1.19 (14.3) | 29/07/2022 | 2.37 (28.4) | 14.1 |
| Czech Republic | 10/03/2020 | 04/01/2022 | 1.82 (21.8) | | | |
| Denmark | 10/03/2020 | 28/01/2022 | 1.89 (22.6) | 03/09/2021 | 1.48 (17.8) | -4.8 |
| England | 21/03/2020 | 28/12/2021 | 1.77 (21.3) | 22/12/2021 | 1.75 (21.1) | -0.2 |
| Finland | 15/03/2020 | 20/02/2022 | 1.94 (23.2) | 31/10/2021 | 1.63 (19.5) | -3.7 |
| France | 13/03/2020 | 24/01/2022 | 1.87 (22.4) | 03/12/2021 | 1.72 (20.7) | -1.7 |
| Germany | 15/03/2020 | 09/03/2022 | 1.98 (23.8) | 18/12/2021 | 1.76 (21.1) | -2.7 |
| Greece | 13/03/2020 | 16/02/2022 | 1.93 (23.2) | 24/01/2022 | 1.87 (22.4) | -0.8 |
| Iceland | 15/03/2020 | 17/02/2022 | 1.93 (23.1) | 25/08/2021 | 1.45 (17.3) | -5.8 |
| Ireland | 14/03/2020 | 03/01/2022 | 1.81 (21.7) | 06/09/2021 | 1.48 (17.8) | -3.9 |
| Israel | 12/03/2020 | 24/01/2022 | 1.87 (22.4) | | | |
| Luxembourg | 12/03/2020 | 27/01/2022 | 1.88 (22.5) | | | |
| New Zealand | 20/03/2020 | 12/04/2022 | 2.06 (24.7) | 01/12/2021 | 1.7 (20.4) | -4.3 |
| Northern Ireland | 20/03/2020 | 09/12/2021 | 1.72 (20.7) | 14/03/2022 | 1.98 (23.8) | 3.1 |
| Paraguay | 09/03/2020 | 20/05/2021 | 1.2 (14.4) | | | |
| Poland | 11/03/2020 | 30/11/2021 | 1.72 (20.7) | | | |
| South Africa | 17/03/2020 | 08/07/2021 | 1.31 (15.7) | | | |
| South Korea | 03/02/2020 | 23/03/2022 | 2.13 (25.6) | 25/10/2021 | 1.72 (20.7) | -4.9 |
| Spain | 09/03/2020 | 09/01/2022 | 1.84 (22) | 31/08/2021 | 1.48 (17.7) | -4.3 |
| Sweden | 15/03/2020 | 12/01/2022 | 1.83 (21.9) | 23/01/2022 | 1.86 (22.3) | 0.4 |
| Switzerland | 15/03/2020 | 27/01/2022 | 1.87 (22.4) | | | |
| The Netherlands | 11/03/2020 | 30/01/2022 | 1.89 (22.7) | | | |
| Wales | 20/03/2020 | 15/11/2021 | 1.66 (19.9) | 24/09/2021 | 1.51 (18.2) | -1.7 |

*Date when 70% of the population has been vaccinated - this was not achieved by all countries within OxCGRT project interval
**Difference calculated as *new duration - interval duration* in months

**Supplementary Table 4**: Pandemic duration calculated when a threshold value of 70.0% of the total population vaccinated had been reached, compared to change point detection using the cumulative COVID-19 case count.

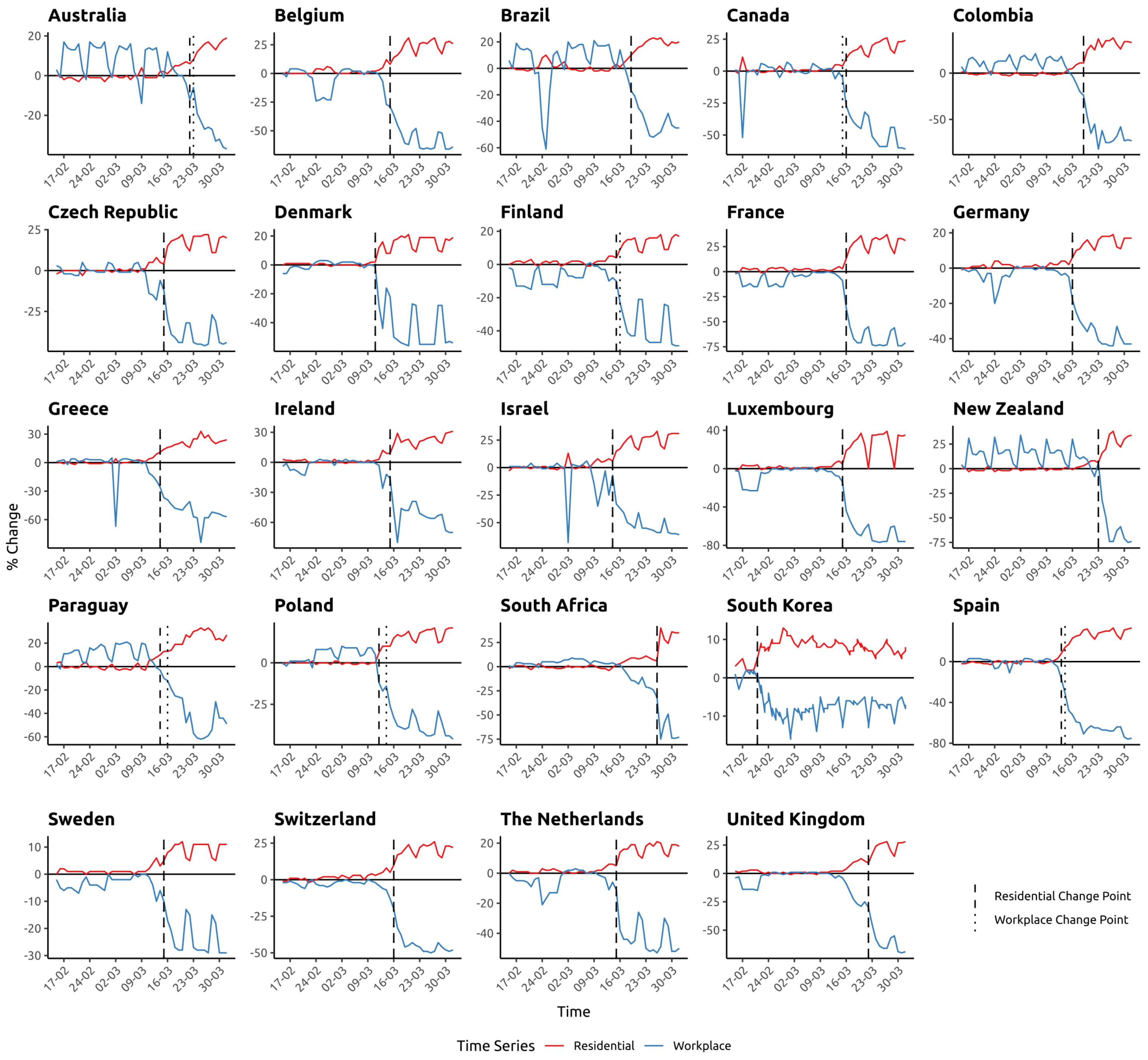


**Supplementary Figure 1**: Containment measure implementation dates determined using change point detection, searching for the most significant change in the mean of the respective time series, applied to workplace and residential time series from the Google COVID-19 Community Mobility Reports (CCMR) data.

| Country | Containment Measure | |
|---|---|---|
| | Implementation | Withdrawal |
| Australia (NSW) | 23/03/2020 | 01/04/2022 |
| Belgium | 15/03/2020 | 16/01/2022 |
| Brazil | 19/03/2020 | 26/05/2021 |
| Canada | 15/03/2020 | 02/01/2022 |
| Colombia | 19/03/2020 | 26/05/2021 |
| Czech Republic | 15/03/2020 | 04/01/2022 |
| Denmark | 11/03/2020 | 28/01/2022 |
| England | 22/03/2020 | 28/12/2021 |
| Finland | 16/03/2020 | 20/02/2022 |
| France | 16/03/2020 | 24/01/2022 |
| Germany | 16/03/2020 | 09/03/2022 |
| Greece | 14/03/2020 | 16/02/2022 |
| Iceland | 15/03/2020 | 17/02/2022 |
| Ireland | 15/03/2020 | 03/01/2022 |
| Israel | 14/03/2020 | 24/01/2022 |
| Luxembourg | 15/03/2020 | 27/01/2022 |
| New Zealand | 23/03/2020 | 12/04/2022 |
| Northern Ireland | 22/03/2020 | 09/12/2021 |
| Paraguay | 16/03/2020 | 20/05/2021 |
| Poland | 14/03/2020 | 30/11/2021 |
| South Africa | 26/03/2020 | 08/07/2021 |
| South Korea | 21/02/2020 | 23/03/2022 |
| Spain | 14/03/2020 | 09/01/2022 |
| Sweden | 15/03/2020 | 12/01/2022 |
| Switzerland | 16/03/2020 | 27/01/2022 |
| The Netherlands | 15/03/2020 | 30/01/2022 |
| Wales | 22/03/2020 | 15/11/2021 |

**Supplementary Table 5**: Final dates used to demarcate pandemic containment measure implementation and withdrawal for each country.

#### 2 Change Point Detection

A simplified explanation of how the At Most One Change (AMOC) algorithm works is detailed below:

1. Data input: the algorithm takes a univariate time series as its data input

2. Potential change points: the algorithm considers every possible location within the data as a potential change point

3. Segment calculation: for each potential change point, the data is divided into two segments: before and after the potential change point

4. Model fitting: for each potential change point, two statistical models are fitted: one for the data before the change point and another for the data after. The fitted model (normal distribution with changing parameters) depends on the function used

5. Parameter estimation: Parameters (eg mean, variance) for each segment are estimated using maximum likelihood estimation

6. Cost function computation: A cost function is computed for each candidate change point (eg sum of squared errors for mean changes)

7. Penalty: a penalty term is added to avoid overfitting. Overfitting occurs when a model is too closely fitted to a limited set of data points. As a result, the model performs well on the data it was trained on but poorly on new, unseen data. This penalty discourages the algorithm from finding too many change points. The penalty function used in this algorithm is the mBIC that penalises models with more parameters, preventing overfitting. The idea is that simpler models are generally preferred unless there's strong evidence to support a more complex one

8. Penalised cost: the total penalised cost is calculated as:

$$\text{Total Cost} = \text{Cost(before)} + \text{Cost(after)} + \text{Penalty}$$

9. Optimal change point: the candidate change point with the smallest penalised cost is selected. If this cost is smaller than the cost of the no-changepoint model (single segment + penalty), a changepoint is declared

#### 3 Population Coverage Adjustment (Simplified)

Three pieces of information obtained for each country and organism:

- Organism case count (detailed in main text)
- Total population
- Percentage of the population captured by the reference laboratory[1]

These data are then used to generate an estimated adjusted case count as follows:

$$\text{Reported Incidence} = \frac{\text{Organism Case Count}}{\text{Total Population} \times \text{Percentage Population Captured}} \qquad \text{(Eq. 1)}$$

$$\text{Adjusted Case Count} = \text{Reported Incidence} \times \text{Total Population} \qquad \text{(Eq. 2)}$$

The 'epiR' package in R (authored by Mark Stevenson) is then used to generate a 95% confidence interval, along with an estimated adjusted incidence rate (see example below).

[1] Assuming the population captured is representative of the general population.

```
# AN EXAMPLE IN R

#install.packages("epiR")
library(epiR)

ncases <- 500
pop_total<-1000000
captured<-0.25
adjusted_case_count<-ncases /captured
npop <- captured*pop_total

tmp <- as.matrix(cbind(ncases, npop))

#ADJUSTED CASE COUNT WITH CONFIDENCE INTERVAL
epi.conf(tmp, ctype = "inc.rate", method = "exact", N = 1000, design = 1,
                                        conf.level = 0.95) * pop_total

#ADJUSTED INCIDENCE RATE WITH CONFIDENCE INTERVAL PER 100,000 POPULATION
epi.conf(tmp, ctype = "inc.rate", method = "exact", N = 1000, design = 1,
         conf.level = 0.95) * 100000
```

### 4 Interrupted Time Series Analysis

#### 4.1 Model Specification

The country-specific models took the following form:

$$\begin{aligned}\ln(\text{adjusted count}_t) =& \beta_0 + \beta_1(T_t) + \beta_2(intervention) \\ &+ \beta_3 \sin\left(\frac{2\pi \cdot M_t}{12}\right) + \beta_4 \cos\left(\frac{2\pi \cdot M_t}{12}\right) \\ &+ \beta_5 \sin\left(\frac{4\pi \cdot M_t}{12}\right) + \beta_6 \cos\left(\frac{4\pi \cdot M_t}{12}\right) \\ &+ \ln(population) + \epsilon\end{aligned} \quad \text{(Eq. 3)}$$

where:

- adjusted count$_t$ represents the value of the cases in month $t$
- $\beta_0$ represents the intercept term
- $\beta_1$ - $\beta_6$ represent the model coefficients
- The $cos$ and $sin$ functions represent the Fourier terms used to control for seasonality
- $T_t$ represents the time in months starting at month 1 (January) in 2018

- $M_t$ represents the month of the year
- ln$(population)$ is the natural logarithmic offset term to account for the population size
- $\epsilon$ is the error term

#### 5 Sensitivity Analyses

| Organism | Model | Pandemic Risk | Withdrawal | Post-Pandemic Risk |
|---|---|---|---|---|
| *Streptococcus pneumoniae* | 1 | 0.41 (0.40-0.42) | 1.48 (1.44-1.52) | 0.91 (0.89-0.93) |
| | 2 | 0.41 (0.40-0.42) | 1.47 (1.42-1.51) | 0.91 (0.89-0.93) |
| | 3 | 0.39 (0.37-0.41) | 1.43 (1.31-1.56) | 0.89 (0.83-0.95) |
| | 4 | 0.37 (0.35-0.39) | 1.37 (1.27-1.48) | 0.86 (0.81-0.93) |
| | 5 | 0.38 (0.36-0.40) | 1.36 (1.26-1.47) | 0.86 (0.80-0.92) |
| *Haemophilus influenzae* | 1 | 0.41 (0.39-0.44) | 1.73 (1.61-1.87) | 1.05 (0.98-1.13) |
| | 2 | 0.41 (0.39-0.43) | 1.75 (1.63-1.88) | 1.05 (0.98-1.13) |
| | 3 | 0.42 (0.38-0.46) | 1.66 (1.54-1.89) | 1.02 (0.90-1.15) |
| | 4 | 0.42 (0.39-0.46) | 1.71 (1.53-1.92) | 1.01 (0.91-1.13) |
| | 5 | 0.41 (0.38-0.45) | 1.71 (1.52-1.92) | 1.01 (0.91-1.13) |
| *Neisseria meningitidis* | 1 | 0.27 (0.25-0.29) | 1.29 (1.13-1.49) | 0.61 (0.55-0.67) |
| | 2 | 0.27 (0.25-0.29) | 1.33 (1.16-1.52) | 0.61 (0.56-0.67) |
| | 3 | 0.26 (0.24-0.29) | 1.14 (0.91-1.43) | 0.60 (0.53-0.67) |
| | 4 | 0.27 (0.24-0.29) | 1.18 (0.97-1.44) | 0.61 (0.54-0.68) |
| | 5 | 0.27 (0.25-0.30) | 1.21 (0.99-1.48) | 0.61 (0.54-0.68) |
| *Streptococcus agalactiae* | 1 | 0.96 (0.88-1.05) | 0.93 (0.84-1.03) | 0.95 (0.85-1.06) |
| | 2 | 0.96 (0.89-1.04) | 0.93 (0.84-1.02) | 0.95 (0.85-1.06) |
| | 3 | 0.97 (0.88-1.07) | 0.93 (0.83-1.03) | 0.95 (0.83-1.08) |
| | 4 | 0.97 (0.88-1.06) | 0.93 (0.84-1.03) | 0.96 (0.85-1.09) |
| | 5 | 0.98 (0.89-1.07) | 0.93 (0.84-1.03) | 0.96 (0.85-1.08) |

Models:

1. Poisson GLM + time-stratified model
2. Poisson GLM + Fourier terms model
3. Quasi-Poisson GLM + time-stratified model
4. Negative binomial GLM + time-stratified model
5. Negative binomial GLM + Fourier terms model

#### 6 Code Availability

A link will appear here.
