## Supplementary figures and images for "Quantifying the impact of the COVID-19 pandemic on invasive bacterial diseases across 27 countries and territories: prospective surveillance by the IRIS Consortium"

### SuppFig_1

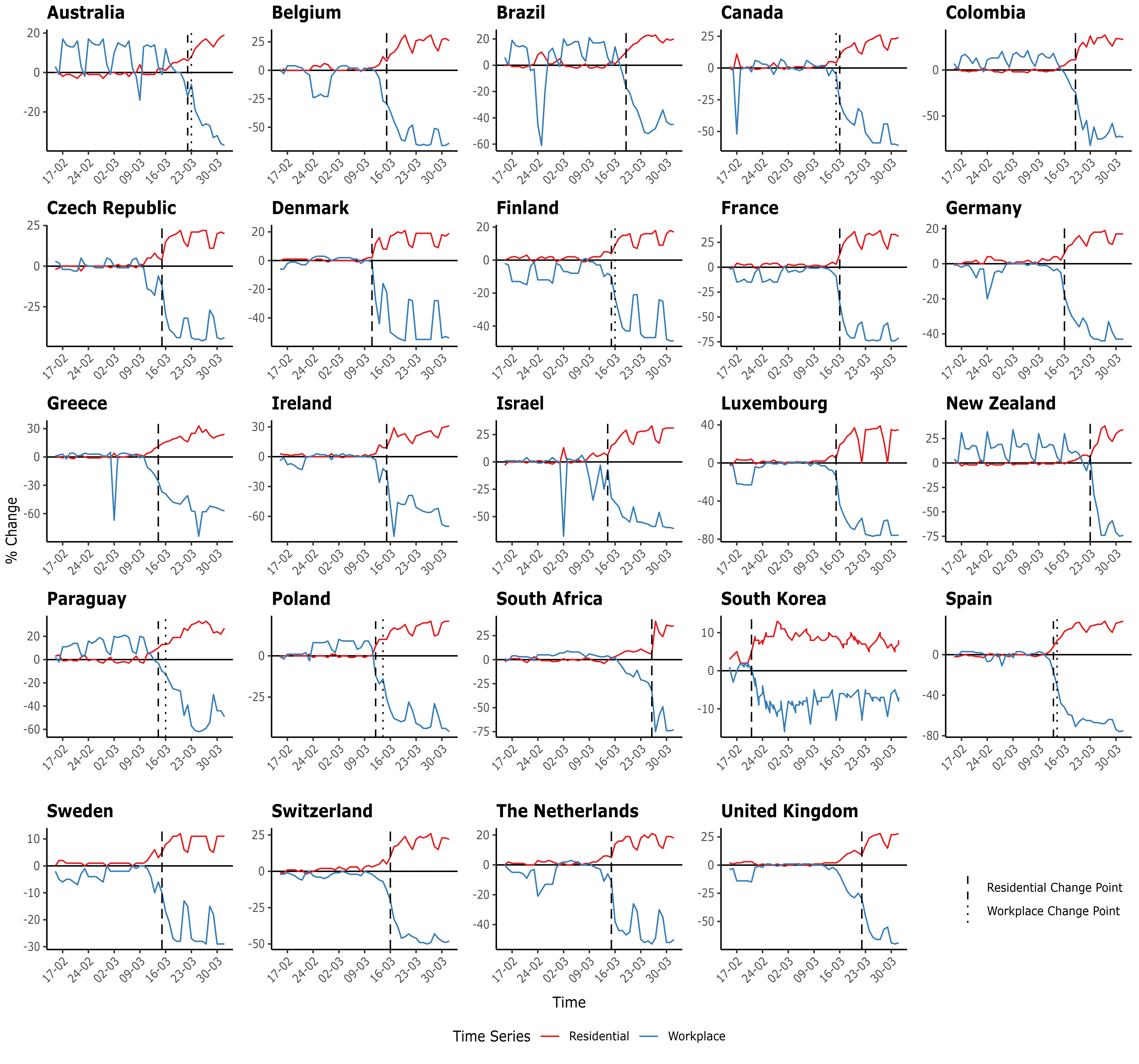
